# Burnout and sleep problems among nurses working in a tertiary hospital in Kathmandu, Nepal

**DOI:** 10.1101/2024.10.09.24315182

**Authors:** Manoj Panthi Kanak, Smriti Pant

## Abstract

Burnout is defined as sustained response to chronic work-related stresses and is often measured in dimensions of emotional exhaustion, disengagement and feeling of low personal accomplishment. Burnout and sleep problems has been closely associated among healthcare providers, confirmed in many observational studies. However, there was a dearth in information regarding burnout and sleep problems among nurses from Nepal, hence this study was conducted. This was a cross-sectional study in which quantitative method was applied. Data was collected from 246 nurses working in the Tribhuvan University Teaching Hospital using simple random sampling method through self-administered questionnaire between March and April, 2022. Structured questions for personal and work-related characteristics were used while Oldenburg Burnout Inventory and Pittsburgh Sleep Quality Index were used to assess burnout and sleep problems, respectively. More than three out of four nurses (78.5%) were found to have burnout, with 88.3% disengaged and 83.0% exhausted. Likewise, almost half of the nurses (40.2%) were found to have a poor sleep quality. Using multivariate logistics regression analysis, factors like position of the nurse (AOR = 6.0, 95% CI; 1.9-18.8) and, slight problem (AOR = 6.6, 95% CI; 3.0-14.7) and somewhat to a big problem (AOR = 6.3, 95% CI; 1.9-20.9) of daytime dysfunction were found to be significantly associated with burnout. Similarly, age, work experience and beds-to-nurses ratio of the department was found to be significantly associated to burnout among the nurses using independent sample t-test. These results indicate the necessity to reduce work stress and manage sleep problems of the nurses prioritizing the nursing staffs occupying junior position. Similarly, establishment of interventions like psychological help desk and support groups for the nurses could be beneficial to mitigate the effects of burnout and sleep problems among nurses.

## Introduction

Nurses are a workforce who have to provide continuous medical care for 24 hours a day in a work environment which is demanding and full of physical and emotional stressors [1,2]. Studies show that stress is a factor that affects nurses on a daily basis and can result in nurses’ absenteeism and aggression as well as reduced productivity and efficiency, diminishing the quality of care and patient safety [3,4]. Shift work has been found to interfere with the circadian and homeostatic regulation of sleep of the nurses causing most nurses working night shifts to struggle adjusting to daytime activities or normal night sleep patterns on their days off [4].

Burnout is defined as a sustained response to chronic work-related stresses and is often measured in three dimensions, namely: a) emotional exhaustion; experience of being emotionally exhausted, b) depersonalization or disengagement; establishment of detached, distant, and cynical relationships with patients and colleagues and, c) feeling of low personal accomplishment and professional failure [5,6]. With increasing work-culture and high demanding jobs, burnout affects workers in a growing number of professions and nurses including other healthcare providers are among the most often affected [5]. Nursing is an occupational area presenting one of the highest levels of work-related stress and burnout, which has been associated to various occupational and personal factors [5,7]. Burnout and sleep problems has been closely associated among healthcare providers and this relation has been confirmed in many observational studies among other types of workers as well [7].

A good sleep is vital for maintaining good health as it affects hormonal levels, mood and physiology. Besides the role of sleep in learning, memory consolidation and motor learning, essential in any professional domain, it has a key role in emotional regulation as well [7]. Complaints of fatigue and insufficient or poor quality of sleep is common among nurses working in 12 hours shifts. Nurses working successively on 12-hours shifts were found to have poor physical and cognitive recovery due to inadequate amount of sleep between the shifts, in a study from the United States [8]. Association between dissatisfaction with sleep patterns and emotional exhaustion among nurses, and a high level of depersonalization among those working the day shift was found in a Brazilian study [1]. Between 57% and 83.2% of shift nurses worldwide report sleep problems, including sleep disturbances, sleep deprivation, and poor sleep quality [4]. Insufficient sleep is a potential cause of burnout, explored in a study among professionals from an information-technology company where less than six hours of sleep at night was the main risk factor for developing burnout [9].

In a multinational study of 2018 which also included Nepal, the level of burnout among nurses working in intensive care units was found to be 52.0%, P = 0.362 [6]. There is no data on prevalence of burnout and sleep problems among nurses in Nepal however, studies among medical students from Nepal has shown prevalence of burnout among 65.9% and poor sleep quality among 38.2% [10,11]. This study was carried out to determine the situation of burnout, sleep quality and, their correlates among nurses working in a tertiary hospital in Kathmandu, Nepal. Data collection was done among 246 nurses working in the Tribhuvan University Teaching Hospital (TUTH) between March and April, 2022.

## Materials and methods

### Study sample

The study followed the simple random sampling method, using computerized number generator without replacement. The details of the study population were obtained from the administrative section of the hospital. A list of the nurses was developed with each unit of these total study population assigned a unique id, and computerized system was used to pick the required sample size from the list. Those selected were noted and contacted through their supervisors, and after receiving their written consent the questionnaire was distributed. The study was carried out in TUTH, Maharajgunj, Nepal. TUTH was selected as study site because TUTH is one of the largest tertiary hospitals in Nepal with a high patient load and with an adequate sample population of nurses.

The sample size was determined using the Cochran’s formula for sample size collection from Sampling Techniques as stated in equation 1 [12].

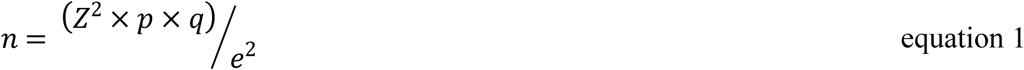

Where

Z= standard normal deviation, usually set at 1.96 which corresponds to 95% confidence level

p = proportion in the target population estimated = 31% [5]

e = degree of accuracy required, usually set at 0.05 level

n = desired sample size which was calculated to be 328

For finite population N = 805 adjusted sample size was calculated to be 233 using equation 2

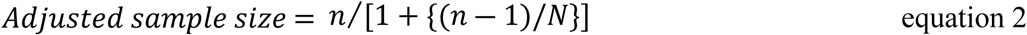

Again, with 10% of non-response rate the final sample size was determined to be 256

Among the 256 self-administered questionnaires distributed, a total of 246 were collected from the respondents indicating a non-response rate of 4%. Questionnaires were distributed in March, 2022 and were collected within April, 2022. Data was entered and processed subsequently after collection.

### Tools for data collection

The study used self-administered questionnaire with three sections; a) socio-demographic and work-related questions, followed by assessment of b) burnout and c) sleep quality. The first section of the questionnaire was used to collect personal and work-related data. It included information regarding the age, gender, education level, marital status, health status, religiosity, cigarette use, alcohol use, exercise, work department and, the number of nurses and beds in the department.

Although Maslach Burnout Inventory is the most commonly used scale to measure burnout, it’s third factor personal accomplishment has been shown to perform weakly [13]. Oldenburg Burnout Inventory (OLBI-S) which contains 16 items questionnaire focusing on the two dimensions of burnout; disengagement and exhaustion was thus used to assess burnout among the nurses in this study [14]. OLBI-S subscales consist of eight items each, among which four are worded positively and four are worded negatively. Participant’s responses were collected on a four-point Likert scale ranging from 1 (strongly agree) to 4 (strongly disagree). Scores from the negatively worded eight items were reversed so that higher score corresponds to higher burnout level.

Pittsburgh Sleep Quality Index (PSQI) was used to assess sleep quality through its seven parameters namely: subjective sleep quality, sleep latency, sleep duration, habitual sleep efficiency, sleep disturbances, use of sleep medication and daytime dysfunction. Each of the seven component has a score between 0 (no problem) to 3 (severe problem) which is added to get the global PSQI score. Both the tools OLBI-S and PSQI have been previously used in Nepal in similar settings [10,11].

### Analytical strategy

The scores from OLBI-S were categorized for prevalence of disengagement and exhaustion using the cutoff of mean ≥ 2.10 from disengagement subscale and mean ≥ 2.25 from exhaustion subscale [15]. Respondents who were found to be both disengaged and exhausted were termed ‘Burnout’ as shown in Figure 1 and the rest were termed ‘No burnout’ for bivariate analysis.

**Figure 1:**
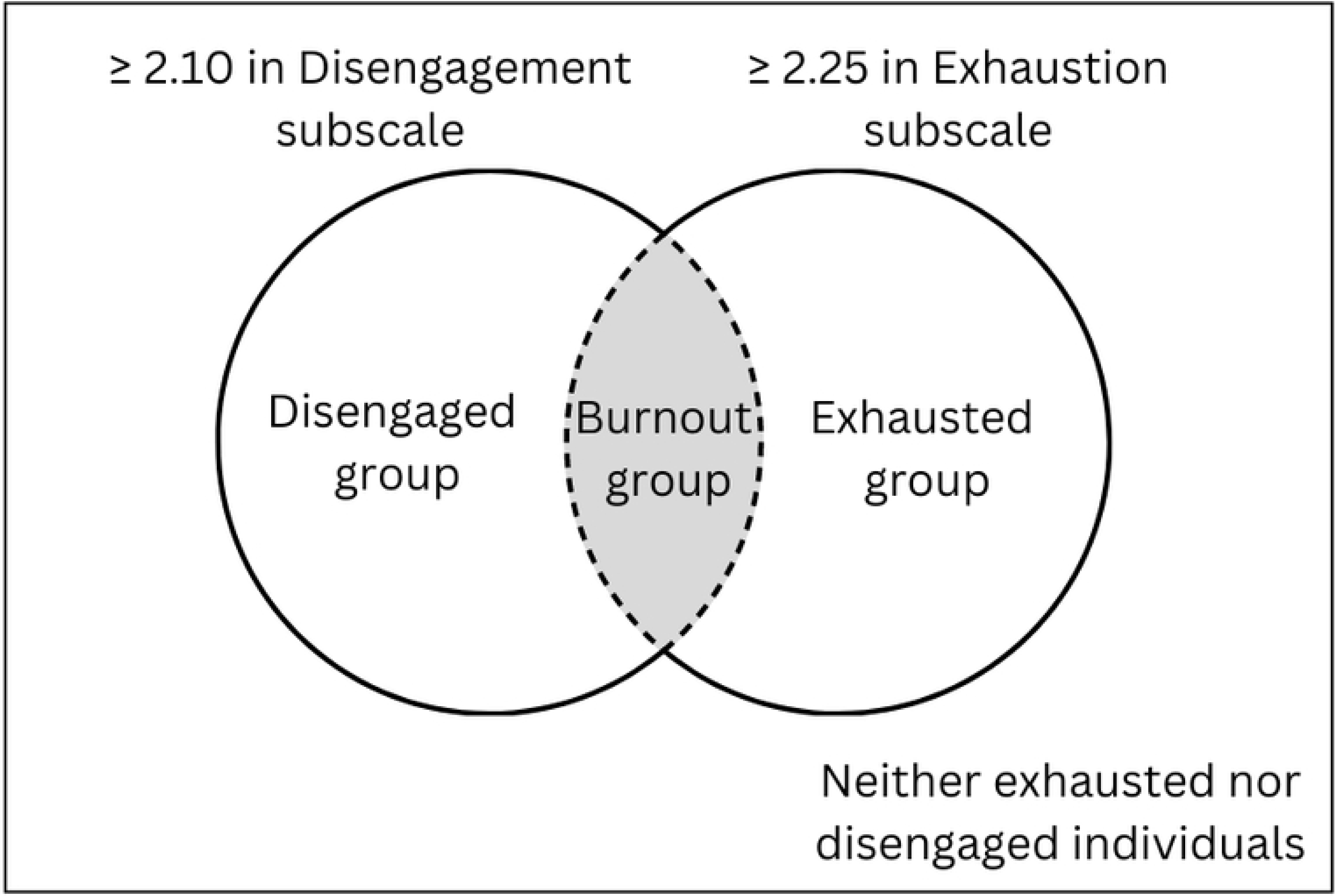
Classification of burnout using OLBI-S.

Higher score in PSQI indicate poorer sleep quality, and a global score of 5 has been established as the cutoff thus, those with score five or more were labelled to have a ‘poor sleep quality’ while the rest were labelled ‘good sleep quality’ [16].

The collected data was entered in EpiData v3.1 and was exported to SPSS 26 for analysis. Descriptive statistics including frequencies and means were calculated from the demographic data, PSQI global scores and Burnout score including sub scores disengagement and exhaustion. Binary logistic regression and independent sample t-test were carried wherever suitable with burnout as an outcome of interest (with 1 = yes and 0 = no). Variables with a p-value less than 0.1 during bivariate analysis were fitted into the multivariate regression model [17]. The adjusted odds ratio was calculated at a 95% CI, and a p-value less than 0.05 was considered statistically significant. The value of Cronbach’s Alpha for 16 items of OLBI-S was 0.787 indicating a good level of internal consistency.

### Ethical consideration

Letter of approval was taken from Institutional Review Committee – Institute of Medicine (Reference number: 339(6-11)E^2^078/079), Central Department of Public Health – Institute of Medicine and TUTH to carry out the research. The research was carried out with the written informed consent of the participants and their voluntary enrollment after thoroughly informing about the research objectives and their responsibilities as research respondents. Confidentiality of the data collected was ensured through coding of each response and the respondents were kept anonymous.

## Results

### Socio-demographic characteristics of the study participants

The age of the 246 study participants was distributed from 20 to 59 years with mean age of 30 years. Of the study participants, 80.5% had a Bachelor degree (Bachelor of Science in Nursing or Bachelor in Nursing), 56.5% were married and 89.4% were religious. In the last month from the data collection, 58.5% had not exercised, 91.9% had not consumed alcohol and 99.2% had not consumed cigarette (Table 1).

**Table 1:** Socio-demographic charactersitics of the study participants.

| Characteristics | Number | Percentage |
| --- | --- | --- |
| Age (years) |  |  |
| Mean | 30 |  |
| SD | ±7.51 |  |
| Highest education |  |  |
| Diploma/PCL Nursing | 37 | 15.0 |
| Bachelors | 198 | 80.5 |
| Masters | 11 | 4.5 |
| Marital status |  |  |
| Married | 139 | 56.5 |
| Unmarried | 106 | 43.1 |
| Divorced | 1 | 0.4 |
| Religious |  |  |
| Yes | 220 | 89.4 |
| No | 26 | 10.6 |
| Exercise |  |  |
| Not during the past month | 114 | 58.5 |
| Less than once a week | 38 | 15.4 |
| Once or twice a week | 36 | 14.6 |
| Three or more times a week | 28 | 11.4 |
| Alcohol consumption |  |  |
| Not during the past month | 226 | 91.9 |
| Less than once a week | 16 | 6.5 |
| Once or twice a week | 3 | 1.2 |
| Three or more times a week | 1 | 0.4 |
| Cigarette consumption |  |  |
| Not during the past month | 244 | 99.2 |
| Less than once a week | 2 | 0.8 |

### Work-related characteristics of the study participants

Among the study participants, 92.7% were working as staff nurse and 46.3% had a work experience of less than 5 years with the mean work experience of 8.20 years. Similarly, 58.12% were stationed at general wards and the mean beds to nurses ratio was 1.4±0.86, which means for every five nurses there were an average of seven beds in the hospital (Table 2).

**Table 2:** Work related characteristics of the study participants.

| Characteristics | Number | Percentage |
| --- | --- | --- |
| Position |  |  |
| Staff Nurse | 228 | 92.7 |
| Nursing Officer | 15 | 6.1 |
| Nursing Supervisor | 3 | 1.2 |
| Work experience (years) |  |  |
| Mean | 8.20 |  |
| SD | $\pm 7.74$ | |
| Department |  |  |
| General wards | 143 | 58.12 |
| Intensive Care Units | 59 | 23.98 |
| Operation Theatre | 16 | 6.50 |
| Emergency | 12 | 4.87 |
| Psychiatry wards | 11 | 4.47 |
| Out Patient Departments | 5 | 2.03 |
| Beds to nurses ratio |  | N=224 |
| Mean | 1.40 |  |
| SD | $\pm 0.86$ | |

### Magnitude of burnout and sleep problems among the study participants

Among the respondents, 88.2% scored ≥2.10 in disengagement subscale and 82.9% scored ≥2.25 in exhaustion subscale of OLBI-S. Respondents who were both disengaged and exhausted i.e., respondents with burnout were found to be 78.5%. Similarly, the mean PSQI global score of the respondents was 5.43 with SD of ±3.035. Among the respondents, ≥ 5 PSQI global score was found among 40.2% implying a poor sleep quality. The mean PSQI global score was higher among respondents who had burnout, which was significantly different to the non-burnout group (p-value = 0.043) ( Table 3).

**Table 3:** Correlation of burnout and sleep quality among the study participants.

| Characteristics | Burnout<br>(%) | No burnout<br>(%) | Total<br>(%) | Chi- square<br>value | p-value |
| --- | --- | --- | --- | --- | --- |
| Overall sleep quality |  |  |  |  |  |
| Mean PSQI ± SD | 5.83±3.085 | 3.98±2.349 | 5.43±3.035 |  | 0.043<sup>#</sup> |
| Overall sleep quality |  |  |  |  |  |
| Good | 106 (72.1) | 41 (27.9) | 147 (59.8) | 8.7 | 0.003 |
| Poor | 87 (87.9) | 12 (12.1) | 99 (40.2) |  |  |
| Subjective sleep quality |  |  |  |  |  |
| Very good | 35 (63.6) | 20 (36.4) | 55 (22.4) | 13.2 | 0.001 |
| Fairly good | 122 (79.7) | 31 (20.3) | 153 (62.2) |  |  |
| Fairly and very bad | 36 (94.7) | 2 (5.3) | 38 (15.4) |  |  |
| Sleep latency |  |  |  |  |  |
| ≤15 minutes | 47 (71.2) | 19 (28.8) | 66 (26.8) | 4.0 | 0.262 |
| 16-30 minutes | 79 (78.2) | 22 (21.8) | 101 (41.1) |  |  |
| 31-60 minutes | 42 (84.0) | 8 (16.0) | 50 (20.3) |  |  |
| >60 minutes | 25 (86.2) | 4 (13.8) | 29 (11.8) |  |  |
| Sleep duration |  |  |  |  |  |
| ≥7 hours | 106 (77.4) | 31 (22.6) | 137 (55.7) | 4.3 | 0.114 |
| 6-7 hours | 60 (75.0) | 20 (25.0) | 80 (32.6) |  |  |
| <6 hours | 27 (93.1) | 2 (6.9) | 29 (11.7) |  |  |
| Sleep efficiency |  |  |  |  |  |
| ≥85% | 119 (78.8) | 32 (21.2) | 151 (61.4) | 2.7 | 0.256 |
| 75-84% | 50 (73.5) | 18 (26.5) | 68 (27.6) |  |  |
| <75% | 24 (88.9) | 3 (11.1) | 27 (11.0) |  |  |
| Sleep disturbance |  |  |  |  |  |
| Not during the past month | 12 (57.1) | 9 (42.9) | 21 (8.5) | 7.4 | 0.024 |
| Less than once a week | 152 (79.2) | 40 (20.8) | 192 (78.1) |  |  |
| Once or more a week | 29 (87.9) | 4 (12.1) | 33 (13.4) |  |  |
| Use of sleep medication |  |  |  |  |  |
| Not during the past month | 179 (78.2) | 50 (21.8) | 229 (93.1) | 0.2 | 0.915 |
| Less than once a week | 9 (81.8) | 2 (18.2) | 11 (4.5) |  |  |
| Once or more a week | 5 (83.3) | 1 (16.7) | 6 (2.4) |  |  |
| Daytime dysfunction |  |  |  |  |  |
| No problem at all | 33 (49.3) | 34 (50.7) | 67 (27.2) | 46.5 | <0.001 |
| Only a very slight problem | 109 (88.6) | 14 (11.4) | 123 (50.0) |  |  |
| Somewhat to a big problem | 51 (91.1) | 5 (8.9) | 56 (22.8) |  |  |

#Independent sample t-test

### Factors associated with burnout

The odds of burnout among study participants who were working in the position of staff nurse were 6.0 (95% CI; 1.9-18.8) times higher than those working in the position of nursing officer or supervisor. In regards to daytime dysfunction, study participants who reported a slight problem were 6.6 (95% CI; 3.0-14.7) times and who reported somewhat to a big problem were 6.3 (95% CI; 1.9-20.9) times more likely to have burnout than those who reported no any problem of day time dysfunction (Table 4). Similarly, using independent sample t-test significant difference was seen in beds-to-nurses ratio (p-value = 0.029) among the respondents with burnout and their counterparts. There also was a significant difference in age (p-value <0.001) and work experience (p-value <0.001) among the respondents with burnout and their counterparts.

**Table 4:** Logistics regression analysis showing association between explanatory variables and burnout among the study participants.

| Explanatory variables | Burnout (%) |  | Crude Odds | Adjusted Odds |
| --- | --- | --- | --- | --- |
|  | Yes | No | Ratio | Ratio |
|  | (n = 193) | (n = 53) | (95% CI) | (95% CI) |
| Position |  |  |  |  |
| Staff nurse | 186 (81.6) | 42 (18.4) | 6.9 (2.5-19.0) | 6.0 (1.9-18.8)* |
| Nursing officer/supervisor | 7 (38.9) | 11 (61.1) | Ref | Ref |
| Subjective sleep quality |  |  |  |  |
| Very good | 35 (63.6) | 20 (36.4) | Ref | Ref |
| Fairly good | 122 (79.7) | 31 (20.3) | 2.5 (1.1-4.4) | 1.3 (0.6-2.8) |
| Fairly and very bad | 36 (94.7) | 2 (5.3) | 10.2 (2.2-47.3) | 3.2 (0.6-17.6) |
| Sleep disturbance |  |  |  |  |
| Not during the past month | 12 (57.1) | 9 (42.9) | Ref | Ref |
| Less than once a week | 152 (79.2) | 40 (20.8) | 2.8 (1.1-7.2) | 1.5 (0.5-4.5) |
| Once or more a week | 29 (87.9) | 4 (12.1) | 5.4 (1.4-21.1) | 1.4 (0.3-7.4) |

| Daytime dysfunction |  |  |  |  |
| --- | --- | --- | --- | --- |
| No problem at all | 33 (49.3) | 34 (50.7) | Ref | Ref |
| Only a very slight problem | 109 (88.6) | 14 (11.4) | 8.0 (3.8-16.7) | 6.6 (3.0-14.7)** |
| Somewhat to a big problem | 51 (91.1) | 5 (8.9) | 10.5 (3.7-29.6) | 6.3 (1.9-20.9)* |

\* p-value less than 0.05
\*\* p-value less than 0.001

## Discussion

This study aimed to find the prevalence of burnout and sleep problems among nurses, and the associated factors of burnout with various socio-demographic and work-related characteristics. The prevalence of burnout among the nurses in this study was 78.5% while, 88.2% were disengaged and 82.9% were exhausted. The prevalence of burnout was found to be much higher than the literature around the globe [1–3,7,18]. Since this is the first study done to map the prevalence of burnout among nurses in Nepal there are no other evidence. However, according to a multinational study that included Nepal the prevalence of burnout among nurses working in intensive care units was 52%, much lower than this study [6]. Similarly, a study among undergraduate medical students in Nepal report prevalence of burnout among 65.9% [10]. Since this study was done post the COVID-19 pandemic the results could have been exacerbated as suggested in a study where pandemic fatigue was found to have a significant negative correlation with mental health. job contentment and sleep quality among the nurses [19]. In this study, prevalence of disengagement was found to be higher than exhaustion which was different to the result from a metanalytic study which reported emotional exhaustion to be the most common dimension of burnout [5].

This study revealed a poor sleep quality among the nurses (40.2%). Although there is no previous data regarding sleep quality among nurses in Nepal, existing evidence suggest poor sleep quality among healthcare providers in a private hospital (48.03%) and undergraduate medical students (38.2%) [11,20]. The mean PSQI score of the nurses in this study was 5.43±3.035, which was lesser in comparison to some studies from around the globe indicating a better sleep quality [2,21–23]. According to a study, nurses have an average sleep of less than 6 hours and this pattern of short sleep could be due to a lack of sleep opportunity rather than to sleep ability [8]. In this study however, the mean sleep hours of the nurses was 6.8±1.1 hours, which is not very less than the recommended seven to eight hours of sleep daily for adults [7].

This study demonstrates a significant association between the position and burnout among the nurses. Other studies also suggest that nurses working in lower positions have greater odds of burnout than their supervisors [1,3,24]. This may be due to greater work load among staff nurses or delegation of work from their supervisors, while lower control over the work. A study suggests that higher level nurses in comparison to their subordinates can better cope high-demanding situations due to greater control over work [25]. Similarly, lack of promotion opportunity and unfair evaluation of work could be a barrier to nurses to their work environment and health, which could be accounted to greater burnout among the staff nurses [26].

In this study, a significant difference in mean PSQI score was seen among nurses with burnout and their counterparts, with higher mean score among burnout group denoting poorer sleep quality. This association between burnout and sleep problem has been revealed in many other studies as well [1,8,27]. Among the seven parameters within sleep quality, daytime dysfunction refers to trouble staying awake and lack of enthusiasm while carrying out daily activities. This study revealed a significant association between burnout and daytime dysfunction among the nurses. Daytime dysfunction could occur due to the demanding nature of shift work causing fatigue in the body and emotional stress among the nurses [1,28].

A significant association was seen between burnout among the nurses and, their age and work experience in this study. Some studies show that younger nurses are at greater risk of burnout, while others reveal that nurses aged over 38–40 years are more vulnerable to burnout. Studies that suggest that longer work experience makes nurses protected to burnout reveal this could be due to familiarization with the scope of work and the work environment [1,3,5]. Beds-to-nurses ratio was found to be significant with burnout among nurses in this study which was congruent to another study. A study suggests that with higher beds-to-nurses ratio there is increase in the workload, and subsequently the work stress of nurses [29]. A difference among emotional exhaustion between nurses working in acute medicine and emergency and accidents departments was noted in another study [5,30]. In this study however, significant association between work department and burnout among the nurses was not observed.

Although many literatures have associated burnout among nurses with higher education i.e., Bachelor or above in comparison to those with Diploma, no such association could be found in this study [6]. Married or living with a spouse and being religious have been linked with protective effect among nurses [3,6]. Being religious is believed to strengthen people when coping with stress and work problems, and it often reduces the negative impact of these on mental health [31]. Likewise, physical exercise has been shown to have a protective effect against burnout and studies report that with greater intensity of physical exercises risk of burnout is lower [32]. However, no such association was found in this study between burnout among the nurses and their marital status, religiousness or physical exercise.

### Conclusion

A very high prevalence of burnout and poor sleep quality was found among the nurses in this study. Position and daytime dysfunction were significant correlates of burnout among the nurses. Similarly, age, work experience and beds-to-nurses ratio of the department was found to be significantly associated to burnout among the nurses. Based on the findings of this study, it is recommended to reduce work stress and manage sleep problems of the nurses especially focusing the junior position nursing staffs. There is a need to reevaluate the burden of work at various departments and ensure staffing of nurses relative to the work burden. Relevant strategies to enhance the mental health of the nurses through interventions like psychological help desk and support groups need to be established. Further prospective studies are required to better understand the cause-effect relationships of risk factors of burnout.

## Data Availability

All data is available without restriction.

https://docs.google.com/spreadsheets/d/1o8st-4gHzoC8fNbLonhqJlMJgUEx69uc/edit?usp=drive_link&ouid=117602478348588754759&rtpof=true&sd=true

## Acknowledgement

We thank all the respondents; the staff nurses, nursing officers and supervisors for their cooperation, valuable participation and honest response during data collection. We are grateful to Tribhuvan University Teaching Hospital for providing administrative approval to conduct the study.

## Supporting information

S1 Data. Data underlying the results of the study

## References

[1] Vidotti V, Ribeiro RP, Galdino MJQ, Martins JT. Burnout Syndrome and shift work among the nursing staff. Rev Lat Am Enfermagem 2018;26. 10.1590/1518-8345.2550.3022.

[2] Dong H, Zhang Q, Sun Z, Sang F, Xu Y. Sleep problems among Chinese clinical nurses working in general hospitals. Occup Med (Chic Ill) 2017;67:534–9. 10.1093/occmed/kqx124.

[3] Lasebikan VO, Oyetunde MO. Burnout among Nurses in a Nigerian General Hospital: Prevalence and Associated Factors. ISRN Nurs 2012;2012:1–6. 10.5402/2012/402157.

[4] Sun Q, Ji X, Zhou W, Liu J. Sleep problems in shift nurses: A brief review and recommendations at both individual and institutional levels. J Nurs Manag 2019. 10.1111/jonm.12656.

[5] Molina-Praena J, Ramirez-Baena L, Gómez-Urquiza JL, Cañadas GR, De la Fuente EI, Cañadas-De la Fuente GA. Levels of burnout and risk factors in medical area nurses: A meta-analytic study. Int J Environ Res Public Health 2018. 10.3390/ijerph15122800.

[6] See KC, Zhao MY, Nakataki E, Chittawatanarat K, Fang W-F, Faruq MO, et al. Professional burnout among physicians and nurses in Asian intensive care units: a multinational survey. Intensive Care Med 2018;44:2079–90. 10.1007/s00134-018-5432-1.

[7] Stewart NH, Arora VM. The Impact of Sleep and Circadian Disorders on Physician Burnout. Chest 2019. 10.1016/j.chest.2019.07.008.

[8] Geiger-Brown J, Rogers VE, Trinkoff AM, Kane RL, Bausell RB, Scharf SM. Sleep, sleepiness, fatigue, and performance of 12-hour-shift nurses. Chronobiol Int 2012. 10.3109/07420528.2011.645752.

[9] Söderström M, Jeding K, Ekstedt M, Perski A, Åkerstedt T. Insufficient sleep predicts clinical burnout. J Occup Health Psychol 2012;17:175–83. 10.1037/a0027518.

[10] Shrestha DB, Katuwal N, Tamang A, Paudel A, Gautam A, Sharma M, et al. Burnout among medical students of a medical college in Kathmandu; A cross-sectional study. PLoS One 2021. 10.1371/journal.pone.0253808.

[11] Paudel K, Adhikari TB, Khanal P, Bhatta R, Paudel R, Bhusal S, et al. Sleep quality and its correlates among undergraduate medical students in Nepal: A cross-sectional study. PLOS Global Public Health 2022;2:e0000012. 10.1371/journal.pgph.0000012.

[12] Cochran WG. Sampling Techniques. 3rd ed. 1977.

[13] Kalliath TJ, O’Driscoll MP, Gillespie DF, Bluedorn AC. A test of the Maslach Burnout Inventory in three samples of healthcare professionals. Work Stress 2000;14:35–50. 10.1080/026783700417212.

[14] Demerouti E, health AB-H of stress and burnout in, 2008 undefined. The Oldenburg Burnout Inventory: A good alternative to measure burnout and engagement. AcademiaEduE Demerouti, AB BakkerHandbook of Stress and Burnout in Health Care, 2008•academiaEdu 2007.

[15] Reis D, Xanthopoulou D, Tsaousis I. Measuring job and academic burnout with the Oldenburg Burnout Inventory (OLBI): Factorial invariance across samples and countries. Burn Res 2015;2:8–18. 10.1016/j.burn.2014.11.001.

[16] Buysse DJ, Reynolds CF, Monk TH, Berman SR, Kupfer DJ. The Pittsburgh Sleep Quality Index: a new instrument for psychiatric practice and research. Psychiatry Res 1989;28:193–213. 10.1016/0165-1781(89)90047-4.

[17] Bursac Z, Gauss CH, Williams DK, Hosmer DW. Purposeful selection of variables in logistic regression. Source Code Biol Med 2008;3:1–8. 10.1186/1751-0473-3-17/TABLES/6.

[18] Rezaei S, Karami Matin B, Hajizadeh M, Soroush A, Nouri B. Prevalence of burnout among nurses in Iran: a systematic review and meta-analysis. Int Nurs Rev 2018;65:361–9. 10.1111/inr.12426.

[19] Labrague LJ. Pandemic fatigue and clinical nurses’ mental health, sleep quality and job contentment during the covid-19 pandemic: The mediating role of resilience. J Nurs Manag 2021;29:1992–2001. 10.1111/jonm.13383.

[20] Kafle B, Tiwari S, Pokhrel A, Shrestha R, Bagale Y, Pahari N. Poor Quality of Sleep among Healthcare Workers in a Tertiary Care Centre. Journal of Nepal Medical Association 2024;62:118–20. 10.31729/jnma.8435.

[21] Demir Zencirci A, Arslan S. Morning-evening type and burnout level as factors influencing sleep quality of shift nurses: a questionnaire study. Croat Med J 2011;52:527–37. 10.3325/cmj.2011.52.527.

[22] Tu Z, He J, Zhou N. Sleep quality and mood symptoms in conscripted frontline nurse in Wuhan, China during COVID-19 outbreak. Medicine 2020;99:e20769. 10.1097/MD.0000000000020769.

[23] Park E, Lee HY, Park CS-Y. Association between sleep quality and nurse productivity among Korean clinical nurses. J Nurs Manag 2018;26:1051–8. 10.1111/jonm.12634.

[24] Ferreira N do N, Lucca SR de. Síndrome de burnout em técnicos de enfermagem de um hospital público do Estado de São Paulo. Revista Brasileira de Epidemiologia 2015;18:68–79. 10.1590/1980-5497201500010006.

[25] Johansson G, Sandahl C, Hasson D. Role stress among first-line nurse managers and registered nurses - a comparative study. J Nurs Manag 2013;21:449–58. 10.1111/j.1365-2834.2011.01311.x.

[26] Thapa DR, Subedi M, Ekström-Bergström A, Areskoug Josefsson K, Krettek A. Facilitators for and barriers to nurses’ work-related health-a qualitative study. BMC Nurs 2022;21:218. 10.1186/s12912-022-01003-z.

[27] Vela-Bueno A, Moreno-Jiménez B, Rodríguez-Muñoz A, Olavarrieta-Bernardino S, Fernández-Mendoza J, De la Cruz-Troca JJ, et al. Insomnia and sleep quality among primary care physicians with low and high burnout levels. J Psychosom Res 2008;64:435–42. 10.1016/j.jpsychores.2007.10.014.

[28] Pereira DS, Araújo TSSL, Gois CFL, Gois Júnior JP, Rodriguez EOL, Santos V dos. Occupational stressors among nurses working in urgent and emergency care units. Rev Gaucha Enferm 2014;35:55–61. 10.1590/1983-1447.2014.01.39824.

[29] Dall’Ora C, Ball J, Reinius M, Griffiths P. Burnout in nursing: a theoretical review. Hum Resour Health 2020;18:1–17. 10.1186/s12960-020-00469-9.

[30] Gillespie M, Melby V. Burnout among nursing staff in accident and emergency and acute medicine: a comparative study. J Clin Nurs 2003;12:842–51. 10.1046/j.1365-2702.2003.00802.x.

[31] Galea M. Assessing the Incremental Validity of Spirituality in Predicting Nurses’ Burnout. Archive for the Psychology of Religion 2014;36:118–36. 10.1163/15736121-12341276.

[32] de Vries JD, Claessens BJC, van Hooff MLM, Geurts SAE, van den Bossche SNJ, Kompier MAJ. Disentangling longitudinal relations between physical activity, work-related fatigue, and task demands. Int Arch Occup Environ Health 2016;89:89–101. 10.1007/s00420-015-1054-x.

